# Sleep Quality and Daytime Sleepiness in Patients with Chronic Obstructive Pulmonary Disease in a Tertiary Care Hospital: An Observational Study

**DOI:** 10.64898/2026.09.17.26363323

**Authors:** Saurav Jha, Aayusha Suwal, Bibek Prasad Sah, Vaibhav Kasaundhan

## Abstract

**INTRODUCTION:** Sleep disturbance is a under-recognized problem among patients with chronic obstructive pulmonary disease and may affect health-related quality of life. Data regarding sleep quality and daytime sleepiness remain limited. This study aimed to determine prevalence of poor sleep quality and daytime sleepiness among patients with chronic obstructive pulmonary disease.

**METHODS:** This cross-sectional study was conducted in patients with chronic obstructive pulmonary disease from March to June 2025. 398 patients were enrolled. Sleep quality and daytime sleepiness were assessed using standardized questionnaires. Data were entered into Microsoft Excel 2016 and analyzed using Statistical Package for the Social Sciences version 20.0. Point estimates at 95% confidence interval were calculated along with frequency and percentage for binary data and mean and standard deviation for continuous data.

**RESULTS:** Among 398 COPD patients, 210 (52.78%; 95% CI, 47.86% to 57.62%) had poor sleep quality (PSQI>5) and 184 (46.23%; 95% CI, 41.39% to 51.14%) had excessive daytime sleepiness (ESS>10). Most participants had moderate-to-very severe COPD (77.89%; 95% CI, 73.56% to 81.69%), with a mean CAT score of 24.36±8.16. Frequent exacerbations (≥2/year) occurred in 63.07% (95% CI, 58.22% to 67.66%) of patients. Sleep latency was prolonged, sleep duration was reduced (mean 5.55±0.80 hours), and daytime dysfunction was common.

**CONCLUSIONS:** Poor sleep quality and excessive daytime sleepiness are highly prevalent among COPD patients and are significantly associated with disease severity and exacerbation frequency. These findings highlight the need for routine sleep screening in COPD care, alongside targeted interventions to improve sleep, symptom control, and quality of life.

## INTRODUCTION

Chronic obstructive pulmonary disease (COPD), a heterogeneous lung condition, is characterized by chronic respiratory symptoms like dyspnea, cough, and expectoration due to persistent airflow limitation resulting from airway and /or alveolar abnormalities.^1^. A community-based study conducted in Nepal reported a COPD prevalence of 8.5% using GOLD criteria.^2^

Sleep disturbance is a common but often under-recognized problem in COPD patients which significantly reduces quality of life.^3^ According to reports, between 50% and 70% of individuals with COPD have sleep-related disorders. Impaired lung functions, nocturnal symptoms, and altered gas exchange during sleep contribute to their poor quality of sleep. Disturbed nighttime sleep frequently results in excessive daytime sleepiness, cognitive impairment, and reduced daily functioning.^3^

Despite the burden of COPD, data on sleep quality and daytime sleepiness remain limited. This study aimed to determine the prevalence of poor sleep quality and excessive daytime sleepiness and the relationship with disease severity and exacerbation frequency.

## METHODS

This was a descriptive cross-sectional study conducted among adult patients diagnosed with COPD at Kathmandu Medical College and teaching hospital from February 2025 to July 2025. The study population consisted of 398 patients with confirmed diagnosis of COPD based on the Global Initiative for Chronic Obstructive Lung Disease (GOLD) criteria. Ethical approval for the study was obtained from the Institutional review board of the Kathmandu Medical College and teaching hospital with reference no. 25122024/07 and written informed consent was obtained from all participants prior to enrollment.

Patients were included if they were adults diagnosed with COPD according to GOLD criteria and willing to participate in the study. Patients with previously diagnosed primary sleep disorders such as obstructive sleep apnea or chronic insomnia, psychiatric illnesses, significant neurological conditions that could confound sleep assessment, those receiving sedatives or hypnotics, and critically ill patients unable to respond to questionnaires were excluded.

The sample size was calculated using Cochran’s formula for descriptive cross-sectional studies.

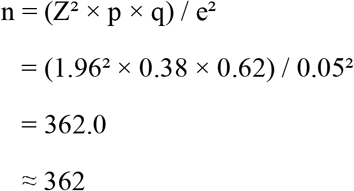

where n is the minimum required sample size, Z is 1.96 at 95% confidence interval, p is the prevalence of sleep-related disorders among patients with COPD taken as 38% based on a previous similar study^4^, q is 1 ™ p, and e is the allowable margin of error set at 5%. Recruitment continued until the planned study end date, during which additional eligible patients were available. Therefore, a total of 398 eligible patients with COPD were included in the final analysis. The larger sample size increased the precision of the estimated prevalence and allowed all eligible participants recruited during the study period to be included.

Disease impact was assessed using the COPD Assessment Test (CAT).^5^ Sleep quality was evaluated using the Pittsburgh Sleep Quality Index (PSQI), a validated questionnaire in which a global score greater than five indicated poor sleep quality.^6^ Daytime sleepiness was assessed using the Epworth Sleepiness Scale (ESS), with a score of ten or more indicating excessive daytime sleepiness.^7^ All questionnaires were administered through face-to-face interviews by trained investigators to ensure consistency and completeness.

Selection bias was reduced by enrolling all eligible participants presenting consecutively during the study period, while information bias was reduced through the use of standardized and validated questionnaires. Data were entered into Microsoft Excel 2016 and analyzed using the Statistical Package for the Social Sciences (SPSS) version 20.0. Continuous variables were expressed as mean and standard deviation, and categorical variables were presented as frequencies and percentages.

## Results

A total of 398 patients with COPD were included in the study. Of these, 331 (83.17%) were males and 67 (16.83%) were females. The mean age of male participants was 62.78±3.88 years, whereas the mean age of female participants was 71.16±1.57 years. The mean body mass index (BMI) of the participants was 24.85 ± 1.52 kg/m^2^, with values ranging from 22.50 to 28.80 kg/m^2^.

According to GOLD staging, mild COPD was found in 22.11% (n=88; 95% CI, 18.31% to 26.44%) of participants, moderate in 36.18% (n=144; 95% CI, 31.61% to 41.01%), severe COPD in 17.59% (n=70; 95% CI, 14.16% to 21.63%), and very severe COPD in 24.12% (n=96; 95% CI, 20.18% to 28.56%), as shown in table 1. Overall, 41.71% (95% CI, 36.97% to 46.61%) of participants had severe to very severe COPD (GOLD stages III–IV), while 58.29% (95% CI, 53.39% to 63.03%) had mild to moderate COPD (GOLD stages I–II).

**Table 1.** Distribution of COPD severity according to GOLD staging (N=398)

| **GOLD Stage** | **N (%)** | **95% CI** |
| --- | --- | --- |
| Mild | 88 (22.11) | 18.31–26.44 |
| Moderate | 144 (36.18) | 31.61–41.01 |
| Severe | 70 (17.59) | 14.16–21.63 |
| Very Severe | 96 (24.12) | 20.18–28.56 |
| **Total** | **398 (100.00)** | — |

The mean CAT score was 24.36±8.16. No participants had a CAT score below 10 (0.00%; 95% CI, 0.00% to 0.96%). 133 (33.41%; 95% CI, 28.96% to 38.19%) had a medium impact (CAT score 10–20), 160 (40.20%; 95% CI, 35.50% to 45.09%) had a high COPD impact (CAT score 21–30), and 105 (26.39%; 95% CI, 22.29% to 30.92%) had a very high impact (CAT score >30). These findings are presented in table 2.

**Table 2.** Distribution of CAT Score Categories in the Study Population (N = 398)

| **CAT Category** | **CAT Score** | **n (%)** | **95% CI** |
| --- | --- | --- | --- |
| Low impact | <10 | 0 (0.00) | 0.00–0.96 |
| Medium impact | 10–20 | 133 (33.41) | 28.96–38.19 |
| High impact | 21–30 | 160 (40.20) | 35.50–45.09 |
| Very high impact | >30 | 105 (26.39) | 22.29–30.92 |

| CAT Category | CAT Score | n (%) | 95% CI |
| --- | --- | --- | --- |
| Total |  | 398 (100.00) | — |

Based on the Pittsburgh Sleep Quality Index (PSQI), 210 (52.78%; 95% CI, 47.86% to 57.62%) participants had poor sleep quality (PSQI > 5), whereas 188 (47.22%; 95% CI, 42.38% to 52.14%) had good sleep quality (PSQI ≤ 5). Regarding sleep latency, 48.74% (n=194; 95% CI, 43.87% to 53.64%) of the participants reported taking 31–60 minutes to fall asleep, while 25.88% (n=103; 95% CI, 21.82% to 30.40%) required more than 60 minutes, 22.61% (n=90; 95% CI, 18.78% to 26.97%) took 16–30 minutes, and only 2.76% (n=11; 95% CI, 1.55% to 4.88%) reported falling asleep within 15 minutes.

Sleep duration was generally short, with 46.48% (n=185; 95% CI, 41.64% to 51.39%) reporting an average of 5 hours of sleep per night, followed by 33.67% (n=134; 95% CI, 29.20% to 38.45%) sleeping 6 hours. Only 13.82% (n=55; 95% CI, 10.77% to 17.56%) reported sleeping 7 hours, whereas 6.03% (n=24; 95% CI, 4.09% to 8.82%) slept 4 hours. The mean sleep duration was 5.55 ± 0.80 hours, with a median of 5.00 hours.

Most participants (64.07%, n=255; 95% CI, 59.24% to 68.63%) reported not using sleep medication during the past month. However, 20.85% (n=83; 95% CI, 17.15% to 25.11%) used sleep medication 1–2 times per week, 9.05% (n=36; 95% CI, 6.61% to 12.27%) used it less than once per week, and 6.03% (n=24; 95% CI, 4.09% to 8.82%) reported using sleep medication three or more times per week.

Regarding subjective sleep quality, over half of the participants (57.29%, n=228; 95% CI, 52.38% to 62.05%) rated their sleep as fairly bad, while 33.92% (n=135; 95% CI, 29.44% to 38.70%) described it as fairly good. Only 2.76% (n=11; 95% CI, 1.55% to 4.88%) reported very good sleep quality, and 6.03% (n=24; 95% CI, 4.09% to 8.82%) reported very bad sleep quality.

With respect to daytime dysfunction, 50.50% (n=201; 95% CI, 45.61% to 55.39%) experienced trouble staying awake less than once per week, while 34.92% (n=139; 95% CI, 30.41% to 39.73%) experienced it 1–2 times per week. Additionally, 9.05% (n=36; 95% CI, 6.61% to 12.27%) reported trouble staying awake three or more times per week, whereas only 5.53% (n=22; 95% CI, 3.68% to 8.23%) did not experience this problem during the past month.

Sleep efficiency was generally moderate among the participants. Nearly half (46.23%, n=184; 95% CI, 41.39% to 51.14%) had a sleep efficiency of 75–84%, while 33.17% (n=132; 95% CI, 28.72% to 37.93%) achieved ≥85% sleep efficiency. Approximately one-fifth (20.60%, n=82; 95% CI, 16.92% to 24.85%) had a sleep efficiency of 65–74% (score 2), and no participants (0.00%; 95% CI, 0.00% to 0.96%) had sleep efficiency below 65%. Table 3 shows all the relevant data analyzed from the PSQI questionnaire.

**Table 3.** Distribution of Sleep Characteristics Among Participants (N=398)

| Variable | Category | n (%) | 95% CI |
| --- | --- | --- | --- |
| Sleep latency (minutes) | ≤15 min | 11 (2.76) | 1.55–4.88 |
|  | 16–30 min | 90 (22.61) | 18.78–26.97 |
|  | 31–60 min | 194 (48.74) | 43.87–53.64 |
|  | >60 min | 103 (25.88) | 21.82–30.40 |
| **Sleep duration (hours)** | 4 hours | 24 (6.03) | 4.09–8.82 |
|  | 5 hours | 185 (46.48) | 41.64–51.39 |
|  | 6 hours | 134 (33.67) | 29.20–38.45 |
|  | 7 hours | 55 (13.82) | 10.77–17.56 |
| **Sleep medication use** | Not in the past month | 255 (64.07) | 59.24–68.63 |
|  | <1 time/week | 36 (9.05) | 6.61–12.27 |
|  | 1–2 times/week | 83 (20.85) | 17.15–25.11 |
|  | ≥3 times/week | 24 (6.03) | 4.09–8.82 |
| **Subjective sleep quality** | Very good | 11 (2.76) | 1.55–4.88 |
|  | Fairly good | 135 (33.92) | 29.44–38.70 |
|  | Fairly bad | 228 (57.29) | 52.38–62.05 |
|  | Very bad | 24 (6.03) | 4.09–8.82 |
| **Sleep efficiency** | ≥85% | 132 (33.17) | 28.72–37.93 |
|  | 75–84% | 184 (46.23) | 41.39–51.14 |
|  | 65–74% | 82 (20.60) | 16.92–24.85 |
|  | <65% | 0 (0.00) | 0.00–0.96 |
| **Trouble staying awake** | Not in the past month | 22 (5.53) | 3.68–8.23 |
|  | <1 time/week | 201 (50.50) | 45.61–55.39 |
|  | 1–2 times/week | 139 (34.92) | 30.41–39.73 |
|  | ≥3 times/week | 36 (9.05) | 6.61–12.27 |

According to the Epworth Sleepiness Scale (ESS), 184 (46.23%; 95% CI, 41.39% to 51.14%) participants exhibited excessive daytime sleepiness (ESS > 10), while 214 (53.77%; 95% CI, 48.86% to 58.61%) had normal daytime sleepiness (ESS ≤ 10). Within these, 8.79% (n=35; 95% CI, 6.39% to 11.98%) had mild excessive daytime sleepiness, 6.03% (n=24; 95% CI, 4.09% to 8.82%) had moderate excessive daytime sleepiness, and 31.41% (n=125; 95% CI, 27.04% to 36.13%) had severe excessive daytime sleepiness. The frequency and percentage of different ESS categories is shown in table 4.

**Table 4.** Distribution of Epworth Sleepiness Scale Scores Among Study Participants (N=398)

| ESS Category | ESS Score Range | n (%) | 95% CI |
| --- | --- | --- | --- |
| Normal daytime sleepiness | 0–10 | 214 (53.77) | 48.86–58.61 |
| Mild excessive daytime sleepiness | 11–12 | 35 (8.79) | 6.39–11.98 |
| Moderate excessive daytime sleepiness | 13–15 | 24 (6.03) | 4.09–8.82 |
| Severe excessive daytime sleepiness | 16–24 | 125 (31.41) | 27.04–36.13 |
| Total |  | 398 (100.00) | — |

251 (63.07%; 95% CI, 58.22% to 67.66%) experienced two or more COPD exacerbations per year, whereas 147 (36.93%; 95% CI, 32.34% to 41.78%) reported fewer than two exacerbations per year. Thus, nearly two-thirds of the study population had a history of frequent exacerbations (≥2 exacerbations annually) as indicated in table 5.

**Table 5.** Frequency of Participants with Two or More COPD Exacerbations per Year(N=398)

| Annual Exacerbations | n (%) | 95% CI |
| --- | --- | --- |
| <2 exacerbations/year | 147 (36.93) | 32.34–41.78 |
| $\geq 2$ exacerbations/year | 251 (63.07) | 58.22–67.66 |
| Total | 398 (100.00) | — |

## DISCUSSION

The principal findings of this study demonstrate that poor sleep quality and increased daytime sleepiness are common among patients with chronic obstructive pulmonary disease (COPD) and are significantly associated with disease severity and exacerbation frequency. In this study, the majority of the patients were male (83.12%) compared to females (16.88%). These findings are similar to the findings of a study conducted in pulmonology clinics of seven Asian cities which demonstrated male predominance (>90%) among COPD patients.^8^ This gender disparity further supports the already established the higher prevalence of risk factors like tobacco smoking and occupational hazards in male population in Asian settings.^9^ Furthermore, COPD in female population is mainly due to biomass exposure from cooking in poorly ventilated rooms, which occur in older women potentially explaining the higher mean age among female population in this study.^9^

In our present study, approximately 78% of the patients had moderate to very severe COPD according to GOLD classification. This finding shows that most patients seeking medical care had moderate to severe airflow limitation (GOLD 2-4), which aligns with the typical presentation of COPD patients seeking medical care.^10^ The mean CAT score in our study was 24.36 ± 8.16, which is comparable with findings reported by Barve et al., who reported a mean score of 23.70 ± 6.28.^11^ Also neither of the study reported low CAT category (<10), which shows that patients presenting in these clinical settings have considerable symptom burdens.^11^ Overall, majority of the patients in both studies had high to very high disease impact (CAT>20) which is 66.6% in this study and 76.6% in the study by Barve et al.^11^

The study found that more than half of the patients had poor sleep quality (PSQI>5) which is consistent with findings of a large population-based analysis from SPIROMICS cohort that reported poor sleep in 51.1% of participants.^12^ Other studies conducted in hospitalized or pulmonary-rehabilitation centers where patients had more advanced disease reported higher prevalence from 73% to as high as 86%.^13,14^ Meanwhile a Saudi Arabian study reported a comparatively lower prevalence of just 32.6%.^15^ These wide variations are likely due to differences in disease severity, study setting, comorbidity burden, and sleep patterns. The prolonged sleep latency observed in this study is in harmony with other COPD cohorts with reported mean sleep latency of 65 minutes.^16^ Vukoja et al. also reported significantly longer sleep latency among patients with COPD suggesting that respiratory impairment contributes directly to difficulty initiating sleep.^4^ This can be explained by persistent load-capacity-drive imbalance produced in COPD patients that worsens in sleep due to decreasing respiratory volumes, altered respiratory muscle tone, and supine positioning that causes nocturnal desaturation and arousal.^17^ Sleep duration was markedly reduced in our study with majority of participants sleeping between 5 to 6 hours only which is far less than the recommended 7-9 hours for healthy adults. Omachi et al. demonstrated that shorter sleep duration in COPD patients is associated with increased respiratory symptoms, poorer health-related quality of life, and a higher frequency of exacerbations.^18^ Nearly one-third of participants reported atleast occasional use of sleep medications which reinforces that sleep problems remain a major contributor to poor quality of life in substantial proportion of COPD patients. This finding is comparable to previous reports suggesting that sleep disturbances in chronic respiratory diseases are often underrecognized and undertreated due to concerns regarding sedative-induced respiratory depression or because sleep complaints are not routinely assessed during clinical encounters.^19^ Subjective sleep quality was poor in participants. Daytime dysfunction was also common, with nearly half of participants reporting difficulty staying awake at least once or twice per week. Night-time respiratory symptoms can disturb sleep and lead to fragmented sleep which over time can lead to excessive daytime fatigue, reduced productivity, and impaired cognitive function.^20^ Also in our study, sleep efficiency was moderately impaired with nearly two-thirds of patient having sleep efficiency below the normal threshold of 85 %. These finding aligns with the previous studies which say that poor sleep efficiency in COPD results from multiple factors including lung hyperinflation, nocturnal oxygen desaturation, and frequent arousals. Polysomnographic studies have further shown that COPD patients experience around 15 arousals per hour on average, considerably more than healthy matched controls which likely contributes to the reduced sleep quality observed in the patients.^21^

In the present study (N = 398), excessive daytime sleepiness (ESS > 10) was present in 46.2% of COPD patients, which is substantially higher than the 17.7% prevalence reported by Stege et al. in stable outpatient COPD populations. Furthermore, while the majority of patients with daytime sleepiness in previous literature presented with mild-to-moderate involvement, our cohort demonstrated a striking predominance of severe excessive daytime sleepiness (ESS 16–24) at 31.4% (n = 125). The high prevalence of severe daytime sleepiness in our study shows significant burden of sleep disruption in COPD patients. It also indicates the presence of nocturnal hypoxemia or co-existing obstructive sleep apnea (overlap syndrome) in this patient population. These findings emphasize the importance of routinely assessing sleep disturbances in patients with COPD to enable timely diagnosis and appropriate management.^22,23^

This study has its own set of limitations. First, depression, anxiety, and other psychological factors that may influence sleep quality in COPD patients were not evaluated. Second, due to the cross-sectional design, causal relationships between COPD severity and sleep disturbances cannot be established. Third, the absence of a non-COPD control group limits the ability to directly compare sleep characteristics with the general population. Nevertheless, the large sample size, rigorous methodology, and comprehensive clinical characterization of COPD patients strengthen the validity of our findings.

Although prior research has highlighted the prevalence and clinical impact of sleep disturbances in COPD, substantial gaps remain in prevention and treatment strategies. Future studies should investigate the specific roles of diverse diagnostic tools and both pharmacologic and non-pharmacologic interventions to better manage sleep disturbances and daytime sleepiness in this population.

Overall, these findings demonstrate the interwoven characteristics of sleep quality, daytime sleepiness, exacerbation frequency, and disease severity. These results prove the pressing need for regular and holistic assessment of sleep quality and daytime sleepiness in COPD patients. This study also advocates the need for targeted interventions to address sleep-related issues in COPD patients in order to improve their health status and overall quality of life.

## CONCLUSION

Poor sleep quality and excessive daytime sleepiness are highly prevalent among COPD patients and are significantly associated with disease severity and exacerbation frequency. Prolonged sleep latency, reduced sleep duration, and impaired sleep efficiency further reflect the interwoven characteristics of sleep quality, daytime sleepiness, exacerbation frequency, and disease severity, highlighting the pressing need for routine sleep screening and targeted interventions to improve sleep, symptom control, and overall quality of life.

## Data Availability

All data produced in the present study are available upon reasonable request to the authors

## Authors’ contribution

**Saurav Jha:** Conceptualization, Methodology, Research Design, Literature review, Data collection, Data Analysis, Statistical Analysis, writing original draft, editing and review; **Aayusha Suwal:** Literature review, Data collection, writing original draft, editing and review; **Bibek Shah:** Literature review, Data collection, Manuscript editing and review; **Vaibhav Kasaundhan:** Methodology, Manuscript editing and review;

